# A Hospital-Based Case-Control Study: Analysis of Risk Factors for Venous Thromboembolism at High Altitudes

**DOI:** 10.1101/2025.06.08.25329227

**Authors:** Xue Sun, Yuting Wu, Xin Liu, Bi Ze, Pema Yangzom, LiPing Du, Pema Lhamo, Chuni Lhamo

**Author notes:** Correspondence: **Yuting Wu**. **Abbreviations:**ALT, alanine aminotransferase; AMI, acute myocardial infarction; APTT, activated partial thromboplastin time; AST, aspartate aminotransferase; DVT, deep vein thrombosis; FDP, fibrin degradation product; HA,high-altitude; PE, pulmonary embolism; VTE, venous thromboembolism.

## Abstract

**Introduction:** The incidence of venous thromboembolism (VTE), a leading cause of morbidity and mortality worldwide, exhibits considerable geographical variability. Although VTE is well- documented in low-altitude settings, the role of high-altitude (HA) environments on the occurrence of VTE remains unclear. Here, we aimed to address this gap by examining the prevalence of VTE at the HAs of the Tibet Autonomous Region, China, offering a unique perspective on how altitude influences VTE risk.

**Methods:** In this retrospective analysis, we reviewed the medical records from the Tibet Autonomous Region People’s Hospital between January 2018 and December 2023. We included 1,084 patients diagnosed with VTE in this quantitative synthesis and comprehensively analysed demographic data, clinical outcomes, and anticoagulant treatment patterns. We used statistical methods, including logistic regression, to identify potential risk factors and explore the relationship between HA and VTE incidence.

**Results:** The prevalence of VTE in the Tibet Autonomous Region People’s Hospital between 2013 and 2023 increased annually from 0.74% to 1.12%. Additionally, VTE was mainly associated with the 18–64-year age group, low haemoglobin levels, and hypertension at HA regions. The occurrence of VTE among residents of HA areas was more common than that reported for lower-altitude populations.

**Conclusion:** This study highlights the critical influence of HA environments on VTE prevalence and the need for tailored healthcare approaches. The unique patterns of anticoagulant therapy after discharge highlight the importance of personalized treatment plans. Overall, our findings contribute to the global understanding of VTE at HA regions and the development of tailored treatment for patients with VTE from HA areas.

## 1 Introduction

Venous thromboembolism (VTE), including deep vein thrombosis (DVT) and pulmonary embolism (PE), represents a substantial public health concern owing to its widespread prevalence (Prabhakar et al., 2019), potential for causing severe complications, and risk of recurrence. Specifically, VTE is the third leading cause of vascular mortality worldwide (Srivastava et al., 2020). Epidemiological data reveal a geographic variation in the incidence of VTE, with estimated rates typically between 1–2 per 1,000 person-years in western countries and less than 1 per 1,000 person-years in eastern countries (Tritschler et al., 2018).

Numerous risk factors, including prolonged immobility (such as during lengthy flights or extended bed rest), surgical interventions, cancer, hormonal therapy, pregnancy, and familial predisposition to blood clots, have been implicated in the development of VTE (Lowe et al., 1999; Dicks et al., 2024). However, the specific contribution of each factor to thrombotic risk and the pathogenetic mechanisms underlying their contributions remain unclear. Lifestyle factors, such as smoking and obesity (Cheng et al., 2013), are also associated with the increased risk of developing VTE. High altitudes (HAs) are associated with a hypercoagulable state (Jones et al., 2022; Broggi et al., 2021), potentially increasing the risk of thrombotic events. An increased prevalence of VTE is associated with populations residing at elevated altitudes (Pelouch et al., 1997; Trunk et al., 2019); some studies have suggested that reduced oxygen availability at high altitudes leads to hypercoagulability (Trunk et al., 2019; Agostoni et al., 2000). However, the effects of HA exposure remain poorly understood, potentially increasing the risk of thrombosis. These inconsistencies highlight the need for real-world data to clarify the impact of HA exposure on VTE and determine whether specific populations, such as older adults or individuals with cardiovascular conditions, are more affected by VTE or more susceptible to developing this condition.

The People’s Hospital of the Tibet Autonomous Region in Lhasa, China, located at an altitude of 3,650 m, serves as the largest and most comprehensive healthcare institution in the region. The residents of this region are genetically adapted to live in hypoxic conditions due to the thin air at HAs. This unique demographic factor can influence the occurrence of a wide range of health issues, including cardiovascular health and blood clotting disorders, both of which are closely related to VTE (Fung et al., 2019; Jha et al., 2018). Considering these aspects, in the present study, we aimed to analyse the prevalence of VTE in HA areas of the Tibetan Autonomous Region by statistically comparing the occurrence and risk factors of VTE between residents of HA and plain areas. Through this comparison, we sought to gain a comprehensive understanding of how HA environments influence the risk of VTE. Such information will facilitate the development of targeted prevention and treatment strategies against these risk factors. Additionally, we aimed to explore VTE treatment and management strategies tailored for HA areas, providing specific clinical guidance. The unique geographical, cultural, and healthcare context of Tibet enhances the relevance of the present study and provides insights not typically available from other regions.

## 2 Methods

### 2.1 Study Design

This retrospective cross-sectional study analysed VTE cases using electronic medical records from the People’s Hospital of the Tibet Autonomous Region, China, from January 1, 2018, to December 31, 2023. The study aimed to describe the prevalence, risk factors, and treatment patterns of VTE in HA populations.

### 2.2 Study Setting and Participants

Setting: The study was conducted at the People’s Hospital of the Tibet Autonomous Region (altitude: 3,650 m; catchment population: ∼3.2 million).

Participants: Inclusion Criteria: Hospitalized patients aged ≥18 years. Primary discharge diagnosis of VTE (ICD-10 codes: I80.2 [deep vein thrombosis], I26.9 [pulmonary embolism]). Patients with concurrent diagnoses (e.g., lower limb venous thrombosis and pulmonary embolism) were categorized hierarchically based on clinical severity. Pulmonary embolism (PE) was prioritized over isolated venous thrombosis, as PE represents a more acute and life-threatening manifestation of VTE. For example, a patient with both lower limb thrombosis and low-risk PE was classified under the Pulmonary Embolism Low Risk category. Patients with multiple PE risk strata (e.g., concurrent low- and intermediate-risk PE) were assigned to the highest applicable risk category.

Exclusion Criteria: (1) Short hospitalization duration: Hospital stay <3 days. (2) Specific thrombotic or vascular diagnoses: Arterial thrombosis (ICD-10: I74.9), Thrombotic microangiopathy (ICD-10: M31.1), Acute cerebral infarction (ICD-10: I63.9), Thrombotic haemorrhoids (ICD-10: K64.8), Thrombotic thrombocytopenic purpura (ICD-10: M31.1), Tethered spinal cord syndrome (ICD-10: Q06.8), Portal vein embolism (ICD-10: I81), Mesenteric venous embolism (ICD-10: K55.0), Other venous thrombosis (ICD-10: I82.90) (3) Systemic complications: Multiple organ failure (ICD-10: R65.2). (4) Data limitations: Missing critical clinical data (e.g., incomplete laboratory results or imaging reports).

Data Source: Structured data were extracted from the Hospital’s Information System Information System (HIS) system, including demographics, laboratory results, comorbidities, and medication records.

### 2.3 Variables

Primary outcome: Incidence of VTE. Secondary outcomes: Clinical improvement, mortality. Covariates included age, sex, and ethnicity, haemoglobin level (g/L), D-dimer (mg/L), fibrin degradation products (FDP, mg/L), and creatinine clearance (CrCL, mL/min) estimated via the Cockcroft-Gault equation; hypertension (defined as systolic blood pressure ≥140 mmHg, diastolic ≥90 mmHg, or documented use of antihypertensive medications); diabetes (fasting glucose ≥7.0 mmol/L or use of glucose-lowering agents), history of VTE; and recent surgical history (within 30 days prior to diagnosis). Covariates: Demographics: age, sex, ethnicity (Han/Tibetan). Laboratory Parameters: D-dimer (mg/L), activated partial thromboplastin time (APTT) (s), and fibrin degradation products (FDP, mg/L).

Comorbidities: Acute myocardial infarction (AMI) (I21.9), ischemic stroke (I63.9), cancer (C00– C97).

Confounders: Adjusted for age, sex, and BMI in regression models.

### 2.4 Data Collection and Quality Control

Data Extraction: Trained researchers used a standardized form to collect variables from the HIS. Missing Data: BMI missing in 12.4% of cases handled via multiple imputation (SPSS MVA module). Laboratory parameters with >20% missingness (e.g., homocysteine) were excluded from multivariate analysis.

Bias Control: Selection Bias: To minimize confounding bias in subgroup analyses (e.g., comparing risk factors across age or sex strata), we performed 1:1 matching based on age and sex within the cohort. This internal matching ensured balanced distributions of these covariates when evaluating associations between altitude-related variables and VTE outcomes. Measurement Bias: ICD-10 codes validated against imaging reports (ultrasound/CT) for VTE confirmation.

### 2.5 Statistical Analysis

Software: SPSS 21.0 (IBM) and GraphPad Prism 9.0. Descriptive Statistics: Continuous variables: Mean ± SD (normal distribution) or median [IQR]. Categorical variables: Frequency (%).

Inferential Statistics: Univariate Analysis: Student’s t-test (continuous), chi-square test (categorical). Multivariate Analysis: Binary logistic regression (enter method) to calculate adjusted odds ratios (aOR) with 95% CI. Sensitivity Analysis: Stratified by age groups (<45 vs. ≥45 years). Threshold: Statistical significance set at p<0.05 (two-tailed).

### 2.6 Ethical Considerations

Approved by the Ethics Committee of the Tibet Autonomous Region People’s Hospital(Approval No.: ME-TBHP-25-KJ-028). Waiver of informed consent granted due to retrospective anonymized data use.

## 2 Results

### 3.1 Annual rate of occurrence of VTE

Table 1 illustrates the annual number of diagnosed thrombosis cases relative to the total number of patients discharged from January 1, 2018 to December 31, 2023. Based on the information collected, the prevalence of VTE in the Tibet Autonomous Region People’s Hospital between 2018 and 2023 was found to have annually increased from 0.62% to 1.07%.

**Table 1.** Diagnosed embolism cases and proportion of total discharged patients.

| Year | Number of patients<br>with VTE (N) | The total number of<br>patients during the same<br>period (N) | proportion (%) |
| --- | --- | --- | --- |
| 2018 | 141 | 22678 | 0.62% |
| 2019 | 162 | 19644 | 0.82% |
| 2020 | 122 | 20433 | 0.60% |
| 2021 | 130 | 20995 | 0.62% |
| 2022 | 165 | 17071 | 0.97% |
| 2023 | 219 | 20383 | 1.07% |

Data reflect full calendar years (January 1, 2018 - December 31, 2023).

### 3.2 Baseline characteristics and comorbidities

A total of 1,084 patients were analysed. The proportion of women with VTEs was marginally higher (52.40%) than that of men with VTE (47.60%). Most patients were within the 18–64-year age group (62.84%), with a notable decline in the number of cases as the age increased. Most patients (73.32%) experienced hospital stays of 14 days or fewer, with the frequency of longer stays diminishing as the duration increased. A substantial proportion of patients exhibited clinical improvement (79.02%). The cure rate was 10.76%, and the mortality rate was comparatively low at 1.81%. Among the 1,084 patients, 63 (5.8%) presented with concurrent lower limb thrombosis and pulmonary embolism. These cases were classified under their respective PE risk categories (e.g., low, intermediate, or high risk) to avoid duplication and prioritize acute clinical management. Lower-limb venous thrombosis was the most prevalent type of thrombosis (41.21%). Various subtypes of PE were observed, with low-risk PE being the predominant subtype (17.04%). These cases were classified under their respective PE risk categories (e.g., low, intermediate, or high risk) to avoid duplication and prioritize acute clinical management. Rarer manifestations included mesenteric venous, portal vein, and intracranial venous sinus thrombosis. The Respiratory Department recorded the highest number of patients with VTE (43.56%), followed by the Hepatobiliary Surgery Department (22.47%). Other departments, such as Neurology, Mountain Disease, Cardiovascular, Obstetrics, and Gynaecology departments, reported a lower prevalence of VTE. The full list of comorbidities and their differences is presented in Table 1. Our findings indicate that in HA areas, the highest prevalence was observed among individuals aged 45–64 years (34.19%) (mean age, 54.87 years; 95% CI, 53.16–55.73).

### 3.3 Test indicators of the patients

Analysis of the patient-related test results indicated that most participants (68.79%) exhibited normal alanine aminotransferase levels, whereas a considerable proportion of these participants (26.56%) presented levels above the upper normal limit, possibly indicating liver damage or inflammation. Most individuals (59.37%) had normal aspartate aminotransferase levels, whereas those of 24.97% of the participants exceeded the upper limit, suggesting potential liver or muscle tissue damage. Approximately half of the patients (44.68%) exhibited CrCL below the normal range; this may potentially be associated with reduced muscle mass or physiological normalcy in certain cases (Gustafsson et al., 1981). Approximately half of the patients (50.47%) showed haemoglobin values within the normal levels. A significant portion (71.90%) of patients exhibited an activated partial thromboplastin time above the upper limit, suggesting the presence of potential bleeding disorders or the effect of anticoagulant therapy. A considerable proportion (86.89%) of patients demonstrated elevated D-dimer levels, indicative of an abnormal clotting process, although elevated D-dimer levels can also occur under various conditions, including infection, inflammation, or postoperative states (Rostami et al., 2020; Kumaran et al., 2024). Furthermore, most patients (66.03%) had fibrin degradation product levels exceeding the reference value, potentially indicating abnormal clotting or fibrinolysis. Approximately one-third (30.94%) of patients had elevated homocysteine levels, which may be linked to vitamin B12 or folate deficiencies and represents a known risk factor for cardiovascular diseases (Mallikarjun et al., 2024). Hypoxic conditions at HAs may increase blood viscosity, thereby increasing the risk of thrombosis (Burns et al., 2018; Hudson et al., 1999).

A comprehensive list of the test parameters is presented in Table 2. After excluding 192 cases of wheelchair use, 159 cases of bed rest, and 134 cases with missing BMI data, statistical analysis was conducted on the remaining 599 medical records with recorded BMI. The analysis indicated a minimum BMI of 16.02, maximum BMI of 41.00, mean BMI of 24.87 (SD 4.72), and median BMI (50% percentile) of 24.39.

**Table 2.** Basic demographic and clinical characteristics and medical history of the study population.

| Characteristics | Reports, No. (%) |
| --- | --- |
| **Patient sex** |  |
| Female | 492(52.40) |
| Male | 447 (47.60) |
| Unknown or missing | 0(0) |
| **Nation** |  |
| Ethnic Han | 72 (7.67) |
| Tibetan | 867 (92.33) |
| **Patient age group (years)** |  |
| <18 | 10 (1.06) |
| 18-44 | 269 (28.65) |
| 45-64 | 321 (34.19) |
| 65-74 | 208 (22.15) |
| 75-84 | 115 (12.25) |
| >85 | 16 (1.70) |
| **Length of stay (days)** |  |
| ≤14 | 681(72.32) |
| 14-30 | 225 (23.96) |
| 30-60 | 28 (2.98) |
| >60 | 5 (0.53) |
| **Disease outcomes** |  |
| Cure | 101 (10.76) |
| Improve | 742 (79.02) |
| Healed | 36 (3.83) |
| Death | 17 (1.81) |
| Other | 43 (4.58) |
| **Year** |  |
| 2018 | 141 (15.02) |
| 2019 | 162 (17.25) |
| 2020 | 122 (12.99) |
| 2021 | 130 (13.84) |
| 2022 | 165 (17.57) |
| 2023 | 219 (23.32) |
| **Discharge diagnosis** |  |
| Venous thrombosis lower limb | 387(41.21%) |
| Pulmonary embolism low risk | 160(17.04%) |
| Pulmonary embolism middle risk | 107(11.40%) |
| Pulmonary embolism high risk | 75(7.99%) |
| Pulmonary embolism middle-low risk | 36(3.83%) |
| Pulmonary embolism middle-high risk | 28(2.98%) |
| Pulmonary embolism unstratified | 146(15.55%) |
| **Department visited** |  |
| Blood Rheumatology ward | 11 (1.17) |
| ICU Intensive Care Unit | 10 (1.06) |
| Mountain disease cardiovascular | 69 (7.35) |
| Respiratory Medicine | 409 (43.56) |
| Internal Medicine | 12 (1.28) |
| Neurology department | 109 (11.61) |
| Nephrology department | 17 (1.81) |
| Gastroenterology department | 2 (0.21) |
| Oncology department | 4 (0.43) |
| Hepatobiliary Surgery | 211 (22.47) |
| Orthopaedic Surgery | 19 (2.02) |
| Urological surgery | 2 (0.21) |
| Neurosurgery | 25 (2.66) |
| Ophthalmology | 1 (0.11) |
| Paediatric | 2 (0.21) |
| Otolaryngology | 2 (0.21) |
| Obstetrics & Gynaecology | 34 (3.62) |

Patients with concurrent VTE subtypes (e.g., DVT and PE) were classified under the highest-priority diagnosis (PE > DVT) based on clinical severity. No overlapping cases were counted in multiple categories. PE classifications supersede isolated DVT in overlapping cases (n=63).

### 3.4 Comorbidities

Table 3 lists various comorbidities, along with the number of cases and their respective percentages in the study population for VTE. The statistical results indicated that at HAs, VTE was mainly associated with hypertension (33.12%), followed by acute myocardial infarction (AMI) or ischemic stroke (12.36%). The chi-square test results indicated no statistically significant association between discharge diagnosis and hypertension (p = 0.480). Patients with VTE frequently experience serious cardiovascular events, such as acute AMI or ischemic stroke, indicative of the interconnection of these conditions (Sławek-Szmyt et al., 2024).

**Table 3.** Distribution of clinical laboratory parameters among participants.

| Indicator | Reference Value | Total Number of Samples | Number and Percentage of Patients Within Normal Range | Number and Percentage of Patients Exceeding Upper Reference Limit | Number and Percentage of Patients Below Lower Reference Limit |
| --- | --- | --- | --- | --- | --- |
| ALT (U/L) | 9–50 | 711 | 485 (68.23) | 187 (26.30) | 39 (5.48) |
| AST (U/L) | 15–40 | 711 | 422 (59.37) | 204 (28.69) | 85 (11.95) |
| eGFR(mL/min/1.73m<sup>2</sup>) | <90 | 711 | 331 (46.60) | 29(4.08) | 351 (49.37) |
| Haemoglobin (g/L) | 130–175 | 820 | 417 (50.84) | 155 (18.90) | 249 (30.24) |
| APTT (S) | 28–44 | 726 | 234 (32.28) | 469 (64.60) | 23(3.17) |
| D-Dimer (mg/L) | <0.5 | 726 | 92 (12.70) | 634 (87.32) | N/A |
| FDP (mg/L) | <5 | 726 | 223 (30.66) | 503 (69.28) | N/A |
| Homocysteine (μmol/L) | 15–20 | 154 | 30 (19.84) | 48 (31.17) | 76(49.35) |

ALT, alanine aminotransferase; APTT, activated partial thromboplastin time; AST, aspartate aminotransferase; eGFR, estimated glomerular filtration rate; FDP, fibrin degradation product

### 3.5 Selection of anticoagulant drugs for hospitalized patients with embolism

Upon diagnosis of VTE, immediate anticoagulation therapy was initiated to prevent clot progression and embolism. Anticoagulant selection is guided by patient-specific factors, including renal function, bleeding risks, preferences, drug interactions, and thrombus characteristics. The primary treatment goal is to stabilize the clot and minimize the risk of embolism, which often requires elevated anticoagulant dosages. Upon hospital admission, VTE risk assessment considers factors such as immobilization, surgical history, and comorbidities to determine appropriate thromboprophylaxis and treatment levels (Lutfi et al., 2024; Prandoni et al., 2007). Most patients received either monotherapy (40.68%) or dual antithrombotic therapy (37.08%), reflecting standard regimens. Acute VTE management typically involves the administration of parenteral anticoagulants, such as unfractionated heparin or low-molecular-weight heparin. Specifically, 53.06% of 441 patients who received monotherapy during hospitalization received sodium heparin or enoxaparin sodium injections.

### 3.6 Discharge with medications

We compiled data regarding the antithrombotic medication status at discharge for 1084 patients with venous thrombosis. The details are provided in Table 4, indicating that most patients were discharged and prescribed oral antithrombotics for continued treatment. Among these, 43.54% of the patients with VTE were treated with rivaroxaban tablets alone after discharge, whereas 32.29% were not prescribed antithrombotic medication.

**Table 4.** Prevalence of comorbidities among patients with VTE.

| Comorbidities | Number of Cases (Percentage) |
| --- | --- |
| Cancer | 75 (7.99) |
| Hypertension | 375 (39.94) |
| Diabetes | 58 (6.18) |
| Heart failure or respiratory failure | 42 (4.47) |
| Acute myocardial infarction or ischemic stroke | 107 (11.40) |
| History of VTE (excluding superficial vein thrombosis) | 68 (7.24) |
| Known thrombophilia | 2 (0.21) |
| Long-term bedridden | 27 (2.88) |
| Recent surgical history | 50 (5.32) |
| None | 135 (14.38) |

VTE, venous thromboembolism

**Table 5.**
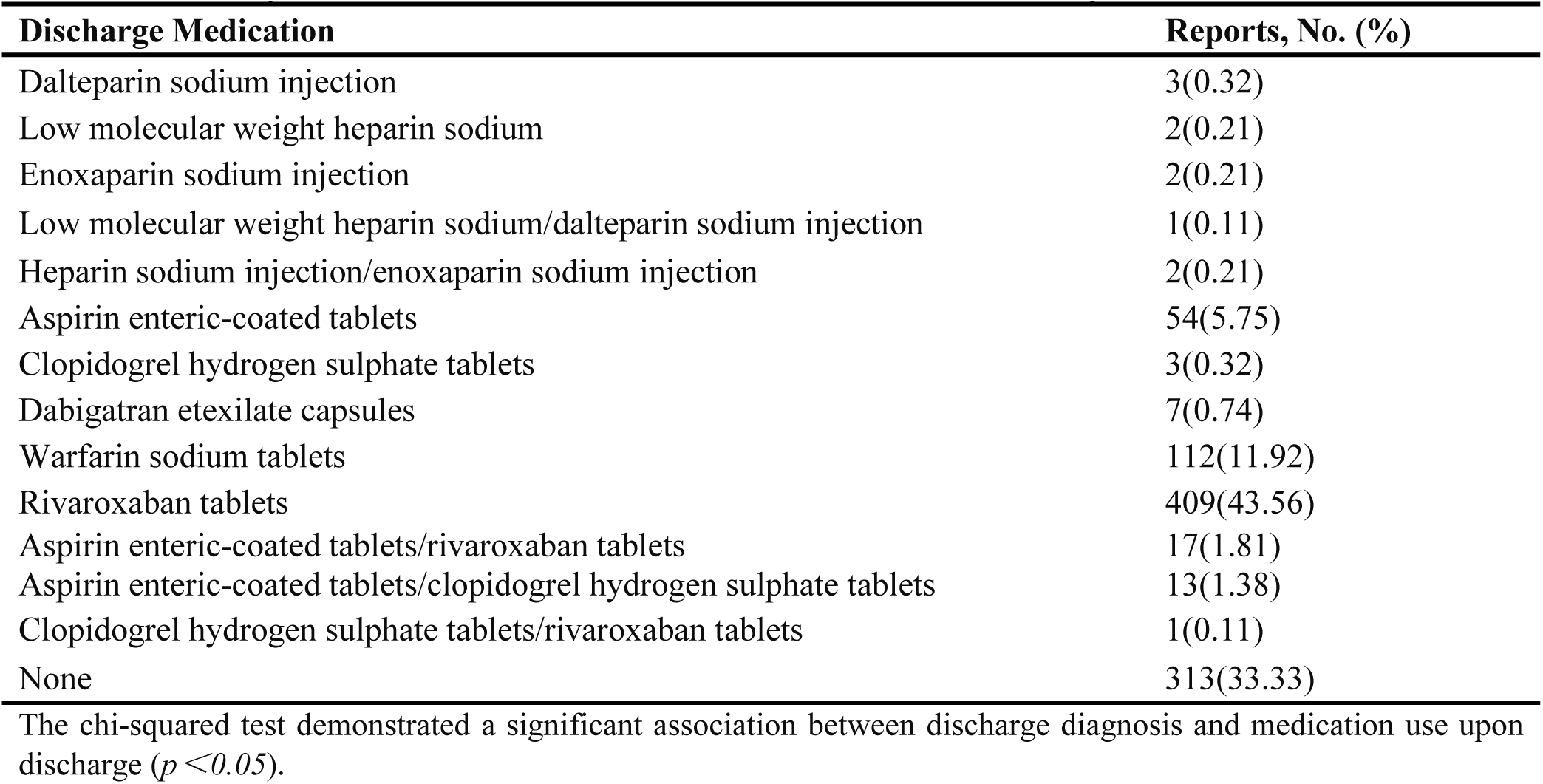
Anticoagulant medications and their case numbers after discharge.

| Discharge Medication | Reports, No. (%) |
| --- | --- |
| Dalteparin sodium injection | 3(0.32) |
| Low molecular weight heparin sodium | 2(0.21) |
| Enoxaparin sodium injection | 2(0.21) |
| Low molecular weight heparin sodium/dalteparin sodium injection | 1(0.11) |
| Heparin sodium injection/enoxaparin sodium injection | 2(0.21) |
| Aspirin enteric-coated tablets | 54(5.75) |
| Clopidogrel hydrogen sulphate tablets | 3(0.32) |
| Dabigatran etexilate capsules | 7(0.74) |
| Warfarin sodium tablets | 112(11.92) |
| Rivaroxaban tablets | 409(43.56) |
| Aspirin enteric-coated tablets/rivaroxaban tablets | 17(1.81) |
| Aspirin enteric-coated tablets/clopidogrel hydrogen sulphate tablets | 13(1.38) |
| Clopidogrel hydrogen sulphate tablets/rivaroxaban tablets | 1(0.11) |
| None | 313(33.33) |

The chi-squared test demonstrated a significant association between discharge diagnosis and medication use upon discharge ( $p < 0.05$ ).

## 4. Discussion

This study exclusively addressed non-cardiogenic venous thrombosis; the implications for AF-related thromboembolism warrant separate investigation. The annual increase in VTE prevalence observed between January 1, 2018, and December 31, 2023, aligns with the rising burden of altitude-related comorbidities in the region, such as hypertension (33.12%) and low haemoglobin levels (30.24%), which were significantly associated with VTE risk in our cohort.

The incidences of DVT and PE significantly increase with age (Patel et al., 2020), particularly after the age of 55. By the age of 80, the incidence rates reach approximately 1 in 100 per year, which is approximately 1000-fold higher than for those aged 45 years or younger. Furthermore, the rates of PE rapidly increase compared to those of DVT in older adult populations (Silverstein et al., 2007). Environmental factors can also lead to heightened DVT and PE incidences in younger demographics. The altered daily activities in HA regions, including occupational and recreational pursuits, may affect blood flow and thrombosis risk. Moreover, the scarcity of medical resources in these areas may impede early diagnosis and treatment of DVT and PE, possibly resulting in a higher incidence among younger individuals.

A notable observation in the present study was that approximately 30.70% of patients with thrombosis had haemoglobin levels below the normal range, suggesting a prevalent incidence of anaemia. This contradicts the common perception that HAs lead to polycythaemia (Gatterer et al., 2024). Approximately half of the patients (49.37%) exhibited reduced eGFR (estimated glomerular filtration rate) below the normal range, suggesting potential renal insufficiency. This finding aligns with previous reports of altered renal haemodynamics in high-altitude populations (Hurtado- Arestegui, A. et al., 2018). Moreover, a significant number of the samples (78.45%) were outside the reference range for homocysteine, with a particularly high percentage (47.51%) falling below the lower limit, indicating potential metabolic anomalies associated with an increased risk of cardiovascular diseases.

The co-occurrence of hypertension was the most prevalent among patients with VTE (33.12 %). Although hypertension is a recognized risk factor for arterial thrombosis (Miraclin et al., 2024), its direct role in VTE risk remains unclear. However, patients with hypertension should be carefully monitored owing to their elevated risk for cardiovascular complications. Following AMI or stroke, immobility, particularly paralysis, inflammation, and hypercoagulability, can predispose patients to VTE (Anderson et al., 2003; Gregson et al., 2019).

In the management of VTE, anticoagulant therapies are fundamental to preventing the formation or enlargement of blood clots. This highlights significant differences in the types and conditions of medications prescribed at discharge, depending on the different discharge diagnoses (Cohen et al., 2020). Rivaroxaban plays a pivotal role in the coagulation cascade. Its benefits include predictable pharmacokinetic profiles, elimination of routine monitoring, and minimal dietary interactions. Rivaroxaban dosing is contingent upon clinical indications and treatment phase, necessitating adjustments for renal function, potential drug interactions, and bleeding risks. For example, a regimen of 15 mg once daily may be appropriate for patients with moderate renal dysfunction after the initial treatment phase. For initial DVT and PE management, 15 mg twice daily with meals is recommended for the first 21 days, followed by a maintenance dose of 20 mg once daily (Gao et al., 2021). In our study, patients administered rivaroxaban typically received conventional treatment regimens, with only one case involving an outpatient medication regimen of rivaroxaban at a dosage of 30 mg once daily and another involving a two-year-old administered a dose of 2 mg once daily for venous sinus thrombosis.

The duration of anticoagulation therapy ranges from a minimum of three months to potentially lifelong treatment, contingent on reversible risk factors, recurrence risk, and the patient’s bleeding risk. One of the key challenges associated with anticoagulant administration in populations at HAs is that standard anticoagulants, such as warfarin, heparin, or direct oral anticoagulant drugs, may display different pharmacokinetics or pharmacodynamics due to the altered vascular environment and reduced oxygen levels. Additionally, given the changes in coagulation function, possibly occurring due to the plateau environment, adjustments in anticoagulant dosing or treatment strategies may be required. These unique environmental factors suggest that standard protocols may not be applicable in HA settings.

However, we acknowledge that the present study has a few limitations. First, relying on data from a single hospital, even one with a significant catchment area, may not fully represent the general population. Second, the lifestyle of Tibetan patients, including dietary habits, physical activity, and cultural practices, could markedly influence the outcomes of anticoagulant therapy. Third, traditional Tibetan medicine, such as herbal treatments or acupuncture, may also interact with conventional anticoagulant therapy. Fourth, genetic predispositions influencing clotting mechanisms in the Tibetan population could introduce bias in conclusions regarding anticoagulant effectiveness. Finally, unaccounted factors, such as climate-related stress and social determinants of health (e.g., social support and education level) may have skewed the analysis, thereby limiting the validity of our findings. Therefore, further research addressing these aspects is necessary to validate our findings and define protocols that are safe and effective for patients under similar conditions.

Further statistical analyses, including those using multivariate models, may help elucidate the relative contributions of these factors to the thrombotic risk in this patient population. Additionally, such analyses could be enhanced by incorporating qualitative data on patient history, medication regimens, and other clinical variables to gain a comprehensive understanding of the pathogenic mechanisms underlying the occurrence of VTE.

Real-world studies provide valuable insights into the impact of HA environments on the prevalence of VTE. These findings can inform the refinement of VTE risk assessment models, incorporating high-altitude environments as a distinct consideration in clinical guidelines. Based on such data, guidelines could recommend deploying more sensitive screening tools or adopting proactive imaging strategies in high-altitude regions, particularly for high-risk groups such as individuals with prolonged immobility, a history of thrombosis, or those recovering from surgery. Furthermore, these studies can help identify populations uniquely vulnerable to high-altitude conditions, such as individuals with polycythaemia, older adults, and postoperative patients, allowing for the development of targeted prevention and treatment strategies. VTE research at HAs also has the potential to garner attention to the health challenges posed by unique geographic conditions globally, paving the way for the establishment of globally standardized guidelines to support patients in similar environments, such as those characterized by hypoxia or extreme climates.

## 5. Conclusion

The main finding of the present cross-sectional study was that the prevalence of VTE in the Tibet Autonomous Region People’s Hospital between 2013 and 2023 increased annually from 0.74% to 1.12%. Additionally, VTE was mainly associated with the 18–64-year age group, low haemoglobin levels, and hypertension at HAs. Our findings provide a better understanding of the effects of HA environments on the occurrence and treatment of VTE, offering more effective prevention and treatment measures for patients with VTE in these regions. Our findings highlight the unique interplay of chronic hypoxia, elevated blood viscosity, and hypertension in driving VTE risk at high altitudes. Hypoxia-induced erythrocytosis, while often adaptive, may paradoxically increase thrombosis susceptibility by altering coagulation dynamics (Jha et al., 2018). These mechanisms, rather than transient external factors, likely underpin the observed trends.

Long-term observations continue to explore the influence of the HA environment on the risk factors for VTE, including hypoxemia, changes in blood viscosity, and lifestyle factors. Examining the differences in VTE incidence between residents of HA and low-altitude areas, along with the underlying physiological mechanisms underlying these differences, can aid the development of targeted strategies for preventing and treating VTE in HA regions. Longitudinal studies and databases should be established for patients with VTE in HA areas to analyse the impact of HA on VTE recurrence and treatment outcomes. Using large-data analytics, in conjunction with genetic, epidemiological, and clinical data, may also elucidate the characteristics and patterns of VTE occurrence in HA regions, providing a foundation for developing prevention strategies and improving treatment efficacy.

## Data Availability

All relevant data are within the manuscript and its Supporting Information files.

## Author Contributions

SX: Conceptualization, Investigation, Methodology, Resources, Validation, Formal Analysis, Visualization, Writing–original draft, Writing–review and editing. WYT: Investigation, Methodology, Resources, Validation, Visualization, Writing–original draft. LX: Investigation, Methodology, Validation. DLP: Investigation, Methodology, Validation, Writing–original draft. ZB: Validation, Writing–review and editing. BMYZ:Investigation, Methodology, Validation. BMLM: Data curation. QNLM: Data curation.

## Conflict of Interest

The authors declare that the research was conducted in the absence of any commercial or financial relationships that could be construed as a potential conflict of interest.

This retrospective study was approved by the Medical Ethics Committee of Tibet Autonomous Region People’s Hospital(Approval No: ME-TBHP-25-KJ-028) with a waiver of informed consent.

## Generative AI statement

The author(s) declare that no Generative AI was used in the creation of this manuscript.

## References

1. Agostoni, P., Cattadori, G., Guazzi, M., Bussotti, M., Conca, C., Lomanto, M., Marenzi, G., & Guazzi, M. D. (2000). Effects of simulated altitude-induced hypoxia on exercise capacity in patients with chronic heart failure. The American journal of medicine, 109(6), 450–455. 10.1016/s0002-9343(00)00532-5

2. Al-Kuraishy, H. M., Al-Gareeb, A. I., Al-Hamash, S. M., Cavalu, S., El-Bouseary, M. M., Sonbol, F. I., & Batiha, G. E. (2022). Changes in the Blood Viscosity in Patients With SARS-CoV-2 Infection. Frontiers in medicine, 9, 876017. 10.3389/fmed.2022.876017

3. Anderson, F. A., Jr, & Spencer, F. A. (2003). Risk factors for venous thromboembolism. Circulation, 107(23 Suppl 1), I9–I16. 10.1161/01.CIR.0000078469.07362.E6

4. Broggi, M. S., Yoon, C. J., Allen, J., Maceroli, M., Moore, T., Schenker, M., & Hernandez-Irizarry, R. (2021). Higher altitude leads to increased risk of venous thromboembolism after acetabular and pelvic ring injury. Journal of clinical orthopaedics and trauma, 19, 192–195. 10.1016/j.jcot.2021.05.026

5. Burns, P., Lipman, G. S., Warner, K., Jurkiewicz, C., Phillips, C., Sanders, L., Soto, M., & Hackett, P. (2019). Altitude Sickness Prevention with Ibuprofen Relative to Acetazolamide. The American journal of medicine, 132(2), 247–251. 10.1016/j.amjmed.2018.10.021

6. Cheng, Y. J., Liu, Z. H., Yao, F. J., Zeng, W. T., Zheng, D. D., Dong, Y. G., & Wu, S. H. (2013). Current and former smoking and risk for venous thromboembolism: a systematic review and meta- analysis. PLoS medicine, 10(9), e1001515. 10.1371/journal.pmed.1001515

7. Cohen, O., Levy-Mendelovich, S., & Ageno, W. (2020). Rivaroxaban for the treatment of venous thromboembolism in pediatric patients. Expert review of cardiovascular therapy, 18(11), 733–741. 10.1080/14779072.2020.1823218

8. Darzi, A. J., Repp, A. B., Spencer, F. A., Morsi, R. Z., Charide, R., Etxeandia-Ikobaltzeta, I., Bauer, K. A., Burnett, A. E., Cushman, M., Dentali, F., Kahn, S. R., Rezende, S. M., Zakai, N. A., Agarwal, A., Karam, S. G., Lotfi, T., Wiercioch, W., Waziry, R., Iorio, A., Akl, E. A., … Schünemann, H. J. (2020). Risk-assessment models for VTE and bleeding in hospitalized medical patients: an overview of systematic reviews. Blood advances, 4(19), 4929–4944. 10.1182/bloodadvances.2020002482

9. Dicks, A. B., Moussallem, E., Stanbro, M., Walls, J., Gandhi, S., & Gray, B. H. (2024). A Comprehensive Review of Risk Factors and Thrombophilia Evaluation in Venous Thromboembolism. Journal of clinical medicine, 13(2), 362. 10.3390/jcm13020362

10. Dix, C., Zeller, J., Stevens, H., Eisenhardt, S. U., Shing, K. S. C. T., Nero, T. L., Morton, C. J., Parker, M. W., Peter, K., & McFadyen, J. D. (2022). C-reactive protein, immunothrombosis and venous thromboembolism. Frontiers in immunology, 13, 1002652. 10.3389/fimmu.2022.1002652

11. Fung, K. P., Chan, K. H., Ng, V., Tsui, P. T., & You, J. H. S. (2019). Health Economic Analysis of Rivaroxaban and Warfarin for Venous Thromboembolism Management in Chinese Patients. Cardiovascular drugs and therapy, 33(3), 331–337. 10.1007/s10557-019-06872-2

12. Gao, Y., & Jin, H. (2021). Rivaroxaban for treatment of livedoid vasculopathy: A systematic review. Dermatologic therapy, 34(5), e15051. 10.1111/dth.15051

13. Gatterer, H., Villafuerte, F. C., Ulrich, S., Bhandari, S. S., Keyes, L. E., & Burtscher, M. (2024). Altitude illnesses. Nature reviews. Disease primers, 10(1), 43. 10.1038/s41572-024-00526-w

14. Gregson, J., Kaptoge, S., Bolton, T., Pennells, L., Willeit, P., Burgess, S., Bell, S., Sweeting, M., Rimm, E. B., Kabrhel, C., Zöller, B., Assmann, G., Gudnason, V., Folsom, A. R., Arndt, V., Fletcher, A., Norman, P. E., Nordestgaard, B. G., Kitamura, A., Mahmoodi, B. K., et al. Emerging Risk Factors Collaboration (2019). Cardiovascular Risk Factors Associated With Venous Thromboembolism. JAMA cardiology, 4(2), 163–173. 10.1001/jamacardio.2018.4537

15. Gustafsson, L., Appelgren, L., & Myrvold, H. E. (1981). Blood flow and in vivo apparent viscosity in working and non-working skeletal muscle of the dog after high and low molecular weight dextran. Circulation research, 48(4), 465–469. 10.1161/01.res.48.4.465

16. Hurtado-Arestegui, A., Plata-Cornejo, R., Cornejo, A., Mas, G., Carbajal, L., Sharma, S., Swenson, E. R., Johnson, R. J., & Pando, J. (2018). Higher prevalence of unrecognized kidney disease at high altitude. Journal of nephrology, 31(2), 263–269. 10.1007/s40620-017-0456-0

17. Hudson, J. G., Bowen, A. L., Navia, P., Rios-Dalenz, J., Pollard, A. J., Williams, D., & Heath, D. (1999). The effect of high altitude on platelet counts, thrombopoietin and erythropoietin levels in young Bolivian airmen visiting the Andes. International journal of biometeorology, 43(2), 85–90. 10.1007/s004840050120

18. Jha, P. K., Sahu, A., Prabhakar, A., Tyagi, T., Chatterjee, T., Arvind, P., Nair, J., Gupta, N., Kumari, B., Nair, V., Bajaj, N., Shanker, J., Sharma, M., Kumar, B., & Ashraf, M. Z. (2018). Genome-Wide Expression Analysis Suggests Hypoxia-Triggered Hyper-Coagulation Leading to Venous Thrombosis at High Altitude. Thrombosis and haemostasis, 118(7), 1279–1295. 10.1055/s-0038-1657770

19. Jones, C. A., Broggi, M. S., Holmes, J. S., Gerlach, E. B., Goedderz, C. J., Ibnamasud, S. H., Hernandez-Irizarry, R., & Schenker, M. L. (2022). High Altitude as a Risk Factor for Venous Thromboembolism in Tibial Plateau Fractures. Cureus, 14(4), e24388. 10.7759/cureus.24388

20. Kumaran, M. S., Singh, S., Mehta, H., & Parsad, D. (2024). Decoding the variability in clinical and laboratory profiles of Chronic Inducible Urticaria vs. Chronic Spontaneous Urticaria - a retrospective study from a tertiary care center. Archives of dermatological research, 316(10), 703. 10.1007/s00403-024-03447-6

21. Lowe, G. D., Haverkate, F., Thompson, S. G., Turner, R. M., Bertina, R. M., Turpie, A. G., & Mannucci, P. M. (1999). Prediction of deep vein thrombosis after elective hip replacement surgery by preoperative clinical and haemostatic variables: the ECAT DVT Study. European Concerted Action on Thrombosis. Thrombosis and haemostasis, 81(6), 879–886.10.1055/s-0037-1614592

22. Lutfi, A., O’Rourke, E., Crowley, M., Craig, E., Worrall, A., Kevane, B., O’Shaughnessy, F., Donnelly, J., Cleary, B., & Áinle, F. N. (2024). VTE risk assessment, prevention and diagnosis in pregnancy. Thrombosis research, 235, 164–174. 10.1016/j.thromres.2024.01.025

23. Miraclin T, A., Bal, D., Sebastian, I., Shanmugasundaram, S., Aaron, S., & Pandian, J. D. (2024). Cerebral Venous Sinus Thrombosis: Current Updates in the Asian Context. Cerebrovascular diseases extra, 14(1), 177–184. 10.1159/000541937

24. Pelouch, V., Kolár, F., Ost’ádal, B., Milerová, M., Cihák, R., & Widimský, J. (1997). Regression of chronic hypoxia-induced pulmonary hypertension, right ventricular hypertrophy, and fibrosis: effect of enalapril. Cardiovascular drugs and therapy, 11(2), 177–185. 10.1023/a:1007788915732

25. Prabhakar, A., Chatterjee, T., Bajaj, N., Tyagi, T., Sahu, A., Gupta, N., Kumari, B., Nair, V., Kumar, B., & Ashraf, M. Z. (2019). Venous thrombosis at altitude presents with distinct biochemical profiles: a comparative study from the Himalayas to the plains. Blood advances, 3(22), 3713–3723. 10.1182/bloodadvances.2018024554

26. Prandoni, P., Noventa, F., Ghirarduzzi, A., Pengo, V., Bernardi, E., Pesavento, R., Iotti, M., Tormene, D., Simioni, P., & Pagnan, A. (2007). The risk of recurrent venous thromboembolism after discontinuing anticoagulation in patients with acute proximal deep vein thrombosis or pulmonary embolism. A prospective cohort study in 1,626 patients. Haematologica, 92(2), 199–205. 10.3324/haematol.10516

27. Rostami, M., & Mansouritorghabeh, H. (2020). D-dimer level in COVID-19 infection: a systematic review. Expert review of hematology, 13(11), 1265–1275. 10.1080/17474086.2020.1831383

28. Shah, N. M., Hussain, S., Cooke, M., O’Hara, J. P., & Mellor, A. (2015). Wilderness medicine at high altitude: recent developments in the field. Open access journal of sports medicine, 6, 319–328. 10.2147/OAJSM.S89856

29. Silverstein, R. L., Bauer, K. A., Cushman, M., Esmon, C. T., Ershler, W. B., & Tracy, R. P. (2007). Venous thrombosis in the elderly: more questions than answers. Blood, 110(9), 3097–3101. 10.1182/blood-2007-06-096545

30. Sławek-Szmyt, S., Araszkiewicz, A., Jankiewicz, S., Grygier, M., Mularek-Kubzdela, T., & Lesiak, M. (2024). Outcomes With Hybrid Catheter-Directed Therapy Compared With Aspiration Thrombectomy for Patients With Intermediate-High Risk Pulmonary Embolism. Cardiovascular drugs and therapy, 10.1007/s10557-024-07562-4. Advance online publication. 10.1007/s10557-024-07562-4

31. Spyropoulos, A. C., Goldin, M., Giannis, D., Diab, W., Wang, J., Khanijo, S., Mignatti, A., Gianos, E., Cohen, M., Sharifova, G., Lund, J. M., Tafur, A., Lewis, P. A., Cohoon, K. P., Rahman, H., Sison, C. P., Lesser, M. L., Ochani, K., Agrawal, N., Hsia, J., … HEP-COVID Investigators (2021). Efficacy and Safety of Therapeutic-Dose Heparin vs Standard Prophylactic or Intermediate-Dose Heparins for Thromboprophylaxis in High-risk Hospitalized Patients With COVID-19: The HEP-COVID Randomized Clinical Trial. JAMA internal medicine, 181(12), 1612–1620. 10.1001/jamainternmed.2021.6203

32. Srivastava, S., Kumari, B., Garg, I., Rai, C., Kumar, V., Yanamandra, U., Singh, J., Panjawani, U., Bansal, A., & Kumar, B. (2020). Targeted gene expression study using TaqMan low density array to gain insights into venous thrombo-embolism (VTE) pathogenesis at high altitude. Blood cells, molecules & diseases, 82, 102421. 10.1016/j.bcmd.2020.102421

33. Steiner, D., & Ay, C. (2022). Risk assessment and primary prevention of VTE in patients with cancer: Advances, challenges, and evidence gaps. Best practice & research. Clinical haematology, 35(1), 101347. 10.1016/j.beha.2022.101347

34. Tritschler, T., Kraaijpoel, N., Le Gal, G., & Wells, P. S. (2018). Venous Thromboembolism: Advances in Diagnosis and Treatment. JAMA, 320(15), 1583–1594. 10.1001/jama.2018.14346

35. Trunk, A. D., Rondina, M. T., & Kaplan, D. A. (2019). Venous Thromboembolism at High Altitude: Our Approach to Patients at Risk. High altitude medicine & biology, 20(4), 331–336. 10.1089/ham.2019.0049

